# COVID-19 breakthrough infection after inactivated vaccine induced robust antibody responses and cross-neutralization of SARS-CoV-2 variants, but less immunity against omicron

**DOI:** 10.1101/2022.01.17.22269415

**Authors:** Nungruthai Suntronwong, Ritthideach Yorsaeng, Jiratchaya Puenpa, Chompoonut Auphimai, Thanunrat Thongmee, Preeyaporn Vichaiwattana, Sitthichai Kanokudom, Thaneeya Duangchinda, Warangkana Chantima, Pattarakul Pakchotanon, Suvichada Assawakosri, Pornjarim Nilyanimit, Sirapa Klinfueng, Lakkhana Wongsrisang, Donchida Srimuan, Thaksaporn Thatsanatorn, Natthinee Sudhinaraset, Nasamon Wanlapakorn, Yong Poovorawan

## Abstract

The emergence of severe acute respiratory syndrome coronavirus 2 (SARS-CoV-2) variants and the waning of immunity in vaccinated individuals is resulting in increased numbers of SARS-CoV-2 breakthrough infections. This study investigated binding antibody responses and neutralizing activities against SARS-CoV-2 variants in patients with COVID-19 who had been fully vaccinated with CoronaVac (*n* = 78), individuals who had been fully vaccinated with CoronaVac but had not contracted COVID-19 (*n* = 170), and individuals who had received AZD1222 as a third vaccination (*n* = 210). Breakthrough infection was generally detected approximately 88 days after the second CoronaVac vaccination (interquartile range 68–100) days. Blood samples were collected at median of 34 days after infection. Binding antibody levels in sera from patients with breakthrough infection were significantly higher than those in individuals who had received AZD1222 as a third vaccination. However, neutralizing activities against wild-type and variants including alpha (B.1.1.7), beta (B.1.351), and delta (B.1.617.2) were comparable in patients with breakthrough infections and individuals who received a third vaccination with AZD1222, which activities are exceeding 90%. Omicron (B.1.1.529) was neutralized less effectively by serum from breakthrough infection patients, with a 6.3-fold reduction compared to delta variants. The study suggests that breakthrough infection after two doses of an inactivated vaccine can induce neutralizing antibody against omicron. Further investigation is needed to assess the long-term persistence of antibody against omicron variant.

---

Since the commencement of the coronavirus disease 2019 (COVID-19) outbreak at the end of 2019, there have been more than 313 million cases of infection [1]. Vaccines are an available tool to prevent and control this threat. Reduced viral susceptibility to vaccine-induced antibodies due to the emergence of severe acute respiratory syndrome coronavirus 2 (SARS-CoV-2) variants, together with the waning of vaccine-induced immunity, is increasing the incidence of COVID-19 breakthrough infection after vaccination [2,3]. Notably however, the study of immunogenicity and neutralization of SARS-CoV-2 variants, particularly omicron (B.1.1.529), is limited [4]. This study aims to determine antibody levels and cross-neutralization in patients with SARS-CoV-2 breakthrough infection following two doses of CoronaVac compared with those in uninfected individuals who received two doses of CoronaVac and those who received AZD1222 as third vaccination.

Patients who had been completely vaccinated with two doses of CoronaVac then subsequently became infected with SARS-CoV-2 as determined via a positive polymerase chain reaction (PCR) test were recruited at the Center of Excellence in Clinical Virology between 21 April and 20 September, 2021. The SARS-CoV-2 variants circulating in Thailand during the study period were alpha (April to June) and delta (July to September) (Figure 1). Controls were unexposed SARS-CoV-2 individuals who had been fully vaccinated with two doses of CoronaVac and screened with anti-nucleocapsid IgG as previously described [5]. Fully vaccinated CoronaVac individuals who received AZD1222 as a third vaccination were also analysed [6].

**Figure 1:**
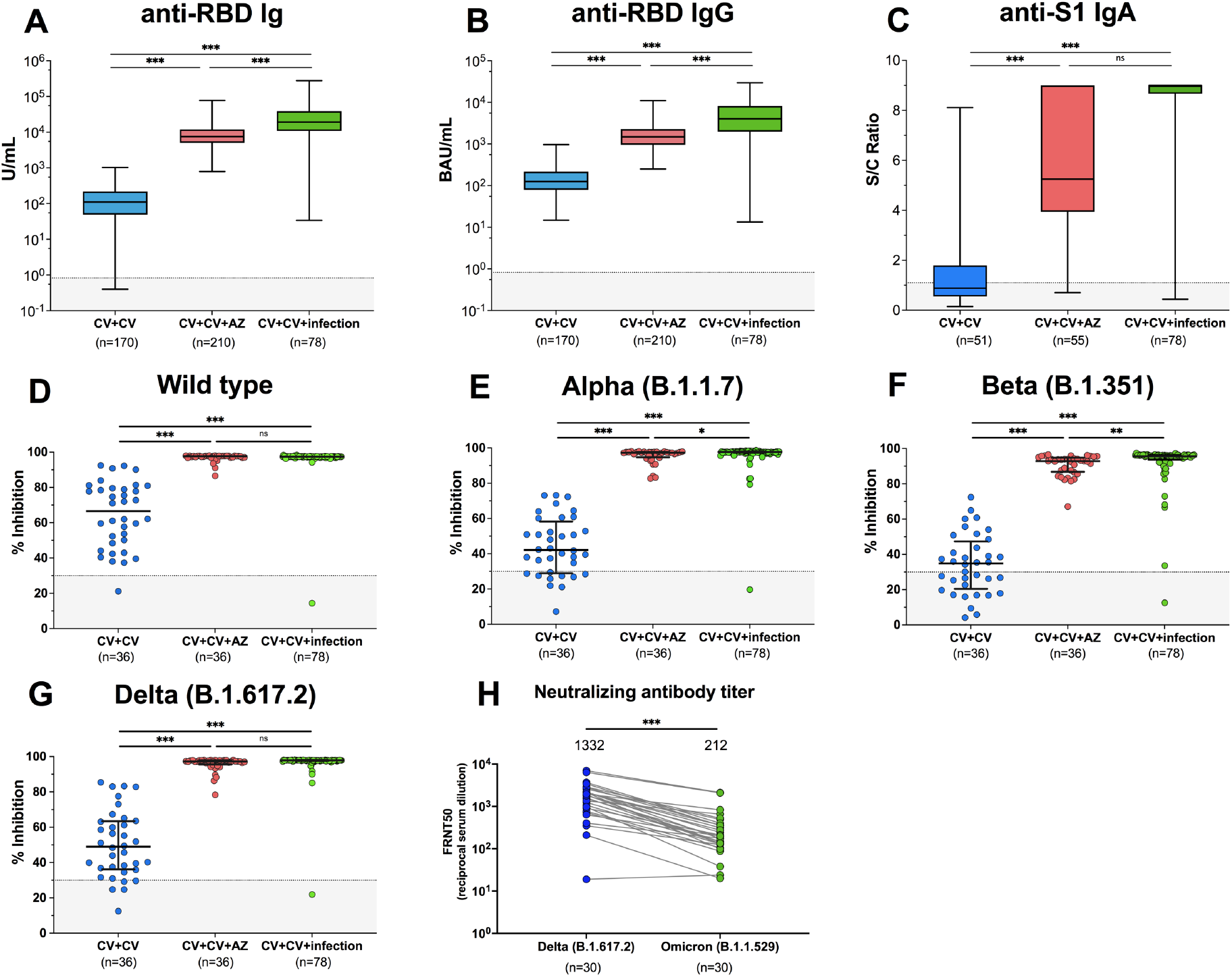
SARS-CoV-2-specific binding antibody responses and neutralizing activities. Immune response of individuals with SARS-CoV-2 breakthrough infection (CV+CV+ Infection) was compared to those with fully vaccinated CoronaVac vaccines without infection (CV+CV) and those who received AZD1222 as third booster (CV+CV+AZ). Serum anti-RBD Ig activity (A), anti-RBD IgG binding antibody units (B), and anti-S1 IgA (optical density of the sample divided by a calibrator; S/C ratio) (C). Neutralizing activities against wild-type (D), B.1.1.7-alpha (E), B.1.351-beta (F), and B.1.617.2-delta (G). Neutralizing antibody titres against B.1.617.2-delta and B.1.1.529-omicron in serum samples from individuals with breakthrough infection determined using focus reduction neutralization test 50 values (H). Geometric mean titres with 95% CIs and median values with IQRs are shown as horizontal bars. Dotted lines indicate cut-off values, and grey shaded areas depict values under the cut-off. Statistics were calculated using Kruskal–Wallis tests with Dunn’s post hoc correction.\**p* < 0.05, \*\**p* < 0.01, \*\*\**p* < 0.001.

All sera were tested for SARS-CoV-2-specific binding antibody responses via enzyme-linked immunosorbent assays, including total immunoglobulin (Ig) against the SARS-CoV-2 receptor-binding domain (RBD), and IgG and IgA specific for SARS-CoV-2 protein. Neutralizing activity against wild-type SARS-CoV-2 and variants including alpha (B.1.1.7; N501Y), beta (B.1.351; K417N, E484K, and N501Y), and delta (B.1.617.2; L452R and T478K) was assessed using a cPass™ SAR-CoV-2 neutralizing antibody detection kit (GenScript, Jiangsu, China) in accordance with the manufacturer’s instructions. Neutralizing antibody titres against delta (B.1.617.2) and omicron (B.1.1.529) were determined using the focus reduction neutralization test as previously described [7].

Geometric mean antibody titres with 95% confidence intervals (CIs) were calculated, and neutralizing activities were expressed as medians and interquartile ranges (IQRs). Differences were assessed using the Kruskal-Wallis method with Dunn’s correction. GraphPad Prism was used to generate figures, and statistics were analysed using SPSS. Statistical significance was defined as *p* < 0.05. The study protocol was conducted in accordance with the Declaration of Helsinki and Good Clinical Practice principles, and it was approved by the Research Ethics Committee of the Faculty of Medicine, Chulalongkorn University (IRB numbers 835/64, 192/64, and 546/64). All participants provided written informed consent.

The group who had been vaccinated with two doses of CoronaVac and subsequently became infected with SARS-CoV-2 contained 78 patients with a mean age of 34.2 years; 58 women (74.4%) and 20 men (25.6%). The median time between the second CoronaVac vaccination and breakthrough infection was 88 days (IQR 68–100 days). Blood samples were collected at 34 days (IQR 29–43) post infection. The group who had been fully vaccinated with CoronaVac but remained unexposed to SARS-CoV-2 infection contained 170 participants with a mean age of 42.3 years; 89 women (52.4%) and 81 men (47.6%). The blood samples were collected at median of 29 days (IQR 27-31) post a second dose vaccination. The group who had been fully vaccinated with CoronaVac then received AZD1222 as a third vaccination contained 210 participants with a mean age of 40 years; 151 women (71.9%) and 59 men (28.1%). The median time between blood sampling and a third dose was 28 days (IQR 20-32) (Supplementary Table 1).

Total RBD-specific Ig was significantly increased in patients with breakthrough infection (18154 units per millilitre (U/mL); 95% CI 13506–24402 U/mL) compared to fully vaccinated individuals without infection (98 U/mL; 95% CI 83–116 U/mL) (Figure 1A). RBD-specific Ig was higher in participants with breakthrough infection than in those who received AZD1222 as a third vaccination (7947 U/mL; 95% CI 7277–8679). Anti-RBD IgG was significantly higher in patients with breakthrough infection (3668 binding antibody units per millilitre (BAU)/mL; 95% CI 2802–4802 BAU/mL) compared to fully vaccinated individuals (128 BAU/mL, 95% CI 114–144 BAU/mL) and those who received AZD1222 as a third vaccination (1492 BAU/mL, 95% CI 1367–1629 BAU/mL) (Figure 1B). Moreover, individuals with breakthrough infection (76/78; 97.4%) and those who received AZD1222 as a third vaccination (54/55; 98.2) were seropositive for anti-S1 IgA. Conversely, only 22/51 (43.1%) of uninfected individuals who had been fully vaccinated with CoronaVac exhibited seropositivity for anti-S1 IgA (Figure 1C). These results indicate that patients with breakthrough infection could mount SARS-CoV-2-specific binding antibody responses.

Among breakthrough infection patients, 77/78 (98.7%) exhibited potential neutralization determined using sVNT against wild-type and SARS-CoV-2 variants. The median neutralizing activities in patients with breakthrough infection were 97.5% for wild-type, 97.7% for alpha, 95.6% for beta, and 97.9% for delta (Supplementary Table 2). Neutralizing activities in breakthrough infection patients were significantly higher than those in unexposed individuals following complete CoronaVac vaccination (*p* < 0.001) (Figure 1D–G). Compared to individuals who received AZD1222 as a third vaccination, in patients with breakthrough infection neutralizing activity was significantly higher for alpha (*p* = 0.02) and beta (*p* < 0.01) variants, but not wild-type or delta variants. This suggests that variant cross-neutralization was improved after breakthrough infection. With respect to neutralizing antibody titres in sera from patients with breakthrough infection determined using live virus neutralization test, FRNT50, the geometric mean titres against delta were 1332 (95% CI 875– 2026), whereas those against omicron were 212 (95% CI 142–316) (Figure 1H). This indicates that omicron is less effectively neutralized by serum antibody derived from breakthrough infection, with a 6.3-fold reduction compared to delta (*p* < 0.001).

Our findings indicate that patients with breakthrough infection exhibited potent antibody responses, with both IgG and IgA titres exceeding those of individuals who received AZD1222 as a third vaccination. Notably the levels of anti-RBD IgG in almost all breakthrough patients were higher than 506 BAU/mL, which was the antibody level associated with 80% effectiveness against symptomatic alpha variant infection in a previous report [8]. Breakthrough infection increased neutralizing activity that cross-inhibited SARS-CoV-2 variants corresponding to wild-type, alpha, beta, and delta. In a previous study a similar result was observed in individuals who had received two mRNA SARS-CoV-2 vaccinations and subsequently became infected [9]. This indicates that natural immunity and vaccine-induced immunity combined result in higher neutralizing potency than vaccination or infection alone [10].

Omicron variants are reportedly less susceptible to neutralization by existing immunity after natural infection [11], and vaccine-induced immunity [12]. Notably, sera from patients with breakthrough infection in the present study also exhibited less neutralizing activity against omicron than against delta. Consistent with a previous study [4] neutralization against omicron was 11.7-fold less than that for wild-type, and 7.9-fold less than that for delta. These results suggest immune escape of omicron in breakthrough infection patients. Reduced neutralizing potency against the omicron variant may be associated with many mutations in the spike protein [13].

The limitations of this study include a small sample size, and the SARS-CoV-2 variants that caused breakthrough infections were not identified. Omicron neutralization assays were not conducted using sera from unexposed individuals who had been fully vaccinated with two doses of CoronaVac or those who received AZD1222 as a third vaccination. Our finding highlights that breakthrough infection after two doses of inactivated vaccines can induce neutralizing antibody against omicron. Further investigation is needed to assess the long-term persistence of antibody against omicron variant.

## Data Availability

All data are available in the report. Additional information can be requested from the corresponding author.

## Acknowledgments

This work was supported by the National Research Council of Thailand, the Health Systems Research Institute, the Center of Excellence in Clinical Virology of Chulalongkorn University, and King Chulalongkorn Memorial Hospital. National Center for Genetic Engineering and Biotechnology (BIOTEC Platform No. P2051613). Nungruthai Suntronwong reports that financial support was also provided by the Second Century Fund Fellowship of Chulalongkorn University.

