## Supplementary Information for "COVID-19 breakthrough infection after inactivated vaccine induced robust antibody responses and cross-neutralization of SARS-CoV-2 variants, but less immunity against omicron"

**Supplementary Figure 1:** Percentage of SARS-CoV-2 variants circulating in Thailand between 21 April 2021 and 20 September 2021.

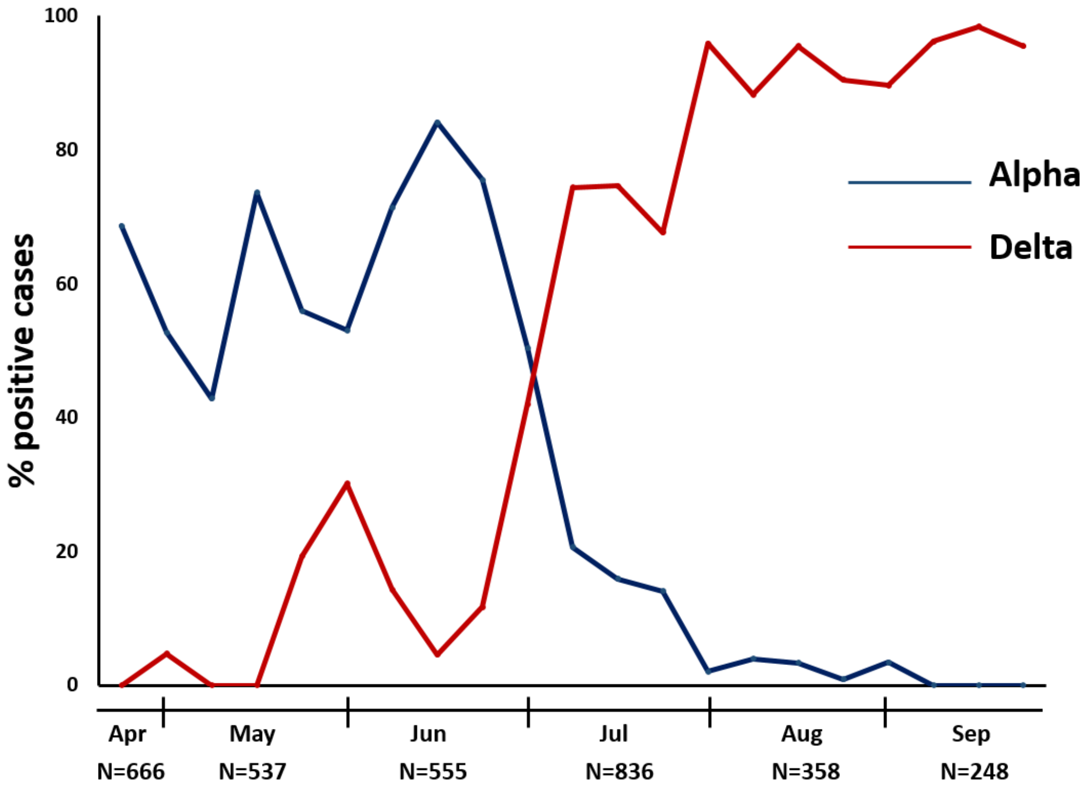


**Supplementary Table 1:** Characteristics of participants in the study.

| **Characteristics** | **CV+CV (*n* = 170)** | **CV+CV+AZ (*n* = 210)** | **CV+CV+INF (*n* = 78)** |
|---|---|---|---|
| Sex (*n*, %) | | | |
| female | 89 (52.4%) | 151 (71.9%) | 58 (74.4%) |
| male | 81 (47.6%) | 59 (28.1%) | 20 (25.6%) |
| Age in years (mean, SD) | 42.3 (9.6) | 40.0 (9.8) | 34.2 (9.9) |
| Interval between 1st and 2nd dose (median, IQR) | 23 (21–26) | 21 (21–26) | 23 (21–28) |
| Interval between 2nd and 3rd dose (median, IQR) | N/A | 70 (61–79) | N/A |

| | | | |
|---|---|---|---|
| Interval between 2nd dose and symptom onset (median, IQR) | N/A | N/A | 88 (68–100) |
| Interval between last vaccination and blood sampling (median, IQR) | 29 (27–31) | 28 (20–32) | N/A |
| Interval between symptom onset and blood sampling (median, IQR) | N/A | N/A | 34 (29–43) |

CV+CV, fully vaccinated with two doses of CoronaVac; CV+CV+AZ, fully vaccinated with two doses of CoronaVac then administered a third vaccination with AZD1222; CV+CV+INF, fully vaccinated with two doses of CoronaVac followed by SARS-CoV-2 breakthrough infection; IQR, interquartile range; N/A, no data available; SD, standard deviation

**Supplementary Table 2:** Binding antibody levels including total anti-RBD Ig, anti-RBD IgG, and anti-S1 IgA. Neutralizing activities of sera from the three groups were measured via surrogate virus neutralization tests.

| | **CV+CV (*n* = 170)** | **CV+CV+AZ (*n* = 210)** | **CV+CV+INF (*n* = 78)** |
|---|---|---|---|
| Anti-RBD-Ig | | | |
| *n* | 170 | 210 | 78 |
| GMT (95% CI) | 98 (83–116) | 7947 (7277–8679) | 18,154 (13,506–24,402) |
| Anti-RBD IgG | | | |
| *n* | 170 | 210 | 78 |
| GMT (95% CI) | 128 (114–144) | 1492 (1367–1629) | 3668 (2802–4802) |
| Anti-S1 IgA | | | |
| *n* | 51 | 55 | 78 |
| Median (IQR) | 0.9 (0.6–1.8) | 5.3 (3.9–9.0) | 9.0 (8.7–9.0) |
| sVNT-wild type | | | |
| *n* | 36 | 36 | 78 |
| Median (IQR) | 66.6 (48.9–79.4) | 97.7 (97.0–97.8) | 97.5 (97.3–97.6) |
| sVNT-B.1.1.7 (alpha) | | | |
| *n* | 36 | 36 | 78 |
| Median (IQR) | 42.1 (29.0–58.3) | 97.2 (94.7–97.7) | 97.7 (97.3–98.0) |
| sVNT-B.1.351 (beta) | | | |

| | | | |
|---|---|---|---|
| *n* | 36 | 36 | 78 |
| Median (IQR) | 34.8 (20.5–47.3) | 92.9 (86.8–94.8) | 95.6 (93.7–96.2) |
| sVNT-B.1.617.2 (delta) | | | |
| *n* | 36 | 36 | 78 |
| Median (IQR) | 48.9 (36.1–63.4) | 97.2 (95.6–97.9) | 97.9 (97.7–98.0) |

CI, confidence interval; CV+CV, fully vaccinated with two doses of CoronaVac; CV+CV+AZ, fully vaccinated with two doses of CoronaVac then administered a third vaccination with AZD1222; CV+CV+INF, fully vaccinated with two doses of CoronaVac followed by SARS-CoV-2 breakthrough infection; GMT, geometric mean titre; IQR, interquartile range; RBD, receptor-binding domain
